# Enteric Pathogens in Soil Adjacent to Public Waste Bins in Maputo, Mozambique Offers Potential for Environmental Surveillance

**DOI:** 10.64898/2026.02.02.26345414

**Authors:** Jack Dalton, Gouthami Rao, Marcia Chiluvane, Victoria Cumbane, David Holcomb, Erin Kowalsky, Amanda Lai, Elly Mataveia, Vanessa Monteiro, Edna Viegas, Joe Brown, Drew Capone

**Author notes:** Corresponding Author: Drew Capone.

## Abstract

Wastewater surveillance has been widely adopted since the COVID-19 pandemic, but non-sewered or onsite sanitation is a common form of sanitation in cities of low- and middle-income countries. Environmental surveillance in these settings requires expanding analyses beyond wastewater. We collected 81 soil samples adjacent to public waste bins inside the sewered and non-sewered areas of Maputo and a 150-meter-wide buffer zone between the two areas, as well as from subsistence farms near the wastewater treatment plant for comparison. We cultured *Escherichia coli* (*E. coli)* using the IDEXX Quanti-Tray/2000 system and determined the prevalence of 29 unique enteric pathogens via RT-qPCR on TaqMan array cards. *E. coli* concentrations were significantly higher (p<.001) in soils adjacent to public waste bins (mean = 5.05×10^5^ per gram) compared to soils from farms (mean = 8.70×10^1^ per gram). The mean number of unique pathogens was higher in soils from the non-sewered area (mean = 7.9, n=32) and the 150-meter buffer area (mean = 10.5, n=10) compared to the sewered area (mean = 4.6, n=20) and soils from farms (mean=3.8, n=19). Findings demonstrate that the presence of enteric pathogens in soils adjacent to public waste bins were associated with neighborhood sanitation infrastructure and may be a useful matrix for surveillance. In high-burden settings with poor sanitation, direct examination of soils and other environmental matrices are potentially scalable means of environmental pathogen surveillance to consider beyond conventional sampling matrices.

## Introduction

Sanitation infrastructure includes onsite systems (e.g., pit latrines, septic tanks) and centralized systems that convey wastewater away from the home for downstream treatment. Piped sewage systems remain a long-term goal in low- and middle-income countries (LMICs), but such systems are not currently financially feasible for many high population density urban neighborhoods [1]. These fecal wastes contain enteric pathogens and failures in the disposal chain may transport waste containing pathogens into the environment. Exposure to a contaminated matrix can occur via well-defined pathways [2], which may result in acute and chronic health outcomes [3].

Wastewater-based epidemiology (WBE) refers to the analysis of fecal wastes for contaminants of interest to infer population health, enabling the assessment of temporal trends, seasonal variability, and scalable monitoring across communities of different sizes and infrastructure[4]. The practice was first introduced in the mid-1900s for monitoring polio and typhoid fever in sewage [5] and since the early 2000s has found renewed interest as a tool for disease surveillance [4, 6, 7]. It has been an important tool for monitoring chemical components, viral targets, SARS-CoV-2, tuberculosis, as well as bacterial and protozoan pathogens [8-10] and been fundamental for global monitoring programs such as the WHO-led Global Polio Surveillance Program [11].

WBE has largely been applied to high resource settings with piped sewer networks because wastewater influent at the treatment plant represents a composite sample of the individuals in the catchment area [12]. Representative sampling in LMICs served by onsite sanitation systems is challenging because there is no matrix analogous to wastewater that provides population level coverage. Limited representative sampling, in addition to limited capacity for ongoing monitoring, partially explains the restricted implementation of WBE in LMICs [13].

Within LMICs, onsite sanitation systems refer to containment technologies such as holding tanks, septic tanks or pit latrines, which require periodic emptying of fecal sludge, which is then transported for treatment at a fecal sludge treatment plant. Centralized treatment refers to wastewater treatment plants, as described above, and fecal sludge treatment plants. Several alternatives to wastewater have been previously investigated to better account for these additional components of the sanitation service chain [14]. Fecal sludge from pit latrines and septic tanks in Maputo, Mozambique contained a mean of 8 unique enteric pathogens and observed pathogen profiles in sludges aligned with the local pediatric infection burden for bacterial and protozoan pathogens [8] [15]. Work in Kenya, Benin, and India suggested that soil could be a suitable matrix for surveillance of soil-transmitted helminth infections [16, 17]. In addition, previous studies have connected solid waste with human pathogen detection in urban environments where open defecation occurs and found solid waste a potential matrix for environmental pathogen surveillance monitoring of SARS-CoV-2 [18, 19].

However, pathogen concentrations in solid waste may be highly heterogenous, given the variable residence time of solid waste in and around public waste bins and frequency of open defecation and urination at these locations. Informal workers commonly sort through solid waste at waste bins and disposal sites looking for recyclables or other valuables that they can sell [20]. The sorting process at waste bins results in large volumes of solid waste being removed from the bin and contacting the ground around the waste bin. It is plausible that fecal contamination in soils adjacent to public waste bins may be more representative of the local burden of disease than the solid waste itself.

The Maputo Sanitation (MapSan) trial was a controlled before-and-after trial to assess the impact of a sanitation intervention (i.e., pour flush to septic tank with soakaway pit for liquid effluent) on stool-based pathogen detection, acute and chronic pediatric health outcomes, and fecal contamination in the environment [21]. At the 24-month follow-up the intervention reduced specific enteric pathogens (i.e., *Trichuris trichiura*, *Shigella flexneri*) in the stool of children who were born after the intervention but had no impact on any pre-specified outcome [21]. The intervention reduced enteric pathogens in soil [22] and antimicrobial resistance genes carried by flies [23], but fecal contamination in the environment remained common 12- and 24-months following the intervention [24].

Previous environmental work in the MapSan trial focused on the domestic domain (i.e., the living environment) [25, 26]. Motivated by the high burden of enteric disease and widespread fecal contamination observed at the 12-month and 24-month trial follow-ups, we decided to investigate fecal contamination in the public domain as part of the 60-month trial follow-up [27, 28]. Our research aim was to determine if the number of unique enteric pathogens in soil adjacent to public waste bins differed between sewered areas, non-sewered areas, and subsistence farms in Maputo, Mozambique. We hypothesized that non-sewered areas would have more unique enteric pathogen detections on average when compared to sewered areas and the subsistence farm plots [29, 30]. Additionally, we aimed to assess the viability of public waste bin soils samples as an additional matrix for environmental pathogen monitoring. This study was not designed to quantify individual-level health risk and is rather an environmental hazard characterization within Maputo.

## Methods

### Sample Collection

We collected 81 soil samples in Maputo, Mozambique using convenience sampling because it was the most time-and-resource efficient approach feasible for this study. No formal sample size calculations were conducted. Maputo is the capital and largest city of Mozambique and has an urban population of over 1.2 million people with nearly 3 million in the greater metropolitan area including both the Matola and Maputo provinces [31]. Soils were collected at the center of the public waste bins located on the pedestrian side of vehicle-trafficked streets within a 3 km radius of Tanzania Avenue and Marien Ngouabi Avenue. These avenues were selected due to the natural division they create within the city between the observed more informal neighborhoods and the colonial-era grid-formatted neighborhoods. We also collected agricultural soil samples from subsistence basin farm plots during transect walks adjacent to the Maputo wastewater treatment plant (WWTP) with the intention to compare these soil samples against the urban public-waste bin soils.

Soil samples were collected from public waste bins in two time periods of February to March 2022 (higher rainfall season) and July to August 2022 (lower rainfall season) to account for potential seasonal rainfall variation. Locations were classified as a non-sewered area, a sewer system area, 150-meter buffer area between the sewered and non-sewered area, and a farm basin area based on existing sanitation infrastructure and land usage. The 150-meter buffer size was determined by a rounded-up distance of two-blocks around the boundary roads between the sewered and non-sewered areas, given that waste bins in this region could receive waste from either the sewered or non-sewered area. This buffer zone represents not only a physical spatial transition, but also a socio behavioral interface where human movement and waste disposal practices from both neighborhood types mix. This buffer zone also represents higher trafficked areas where movement between both neighborhood types occur. The basin farm locations were chosen due to their proximity to the wastewater treatment plant. Samples were additionally categorized by sun exposure (sun, partial sun, shade), and soil conditions (visibly wet, dry) at the time of sampling.

We collected soil samples at the center of a public waste bin, one meter from the edge of the bin, on the pedestrian side opposite of the street. A 10x10x1cm volume of soil was homogenized with a sterile lab scoopula and transferred into three 2ml cryovials and one 5ml cyrovial. Soil was stored in a cooler with ice packs at 4°C and transported to the *Centro de Investigação e Treino em Saúde da Polana Caniço* (CISPOC) laboratory in Maputo, Mozambique within four hours of collection.

### Culture and Molecular Analysis

We cultured *E. coli* from soil using the IDEXX Quanti-Tray/2000 system. One gram of soil (wet weight) was added to 100 mL of autoclaved distilled water and manually shaken for 2 minutes [24]. Five grams of soil was dried using the microwave oven method to determine moisture content [32]. Field blanks were not collected, but a negative control was run each day of lab analysis. Soil sample cryovials were shipped on dry ice (-80°C) with temperature monitoring to the University of North Carolina at Chapel Hill, North Carolina for molecular analysis.

RNA and DNA were extracted from 1.5 g of individual soil samples using the Qiagen RNeasy PowerSoil Total RNA kits and then with the Qiagen RNeasy PowerSoil DNA Elution Kit. Bovine Herpes Virus (*BHV*) and Bovine Respiratory Syncytial Virus (*BRSV*) were spiked into each sample as extraction positive controls for DNA and RNA, respectively. Methodology for the rest of sample preparation and extraction followed the manufacturer’s protocol. Post sample extraction, DNA and total RNA extracts were combined and then stored at – 80°C freezer.

Combined extracts were thawed once and analyzed for the presence of enteric pathogens, fecal source tracking markers, and the integron integrase gene (i.e., marker of anthropogenic antimicrobial resistance) using a TaqMan Array Card (TAC) on an Applied Biosystems QuantStudio 7 Flex. 10% were of samples run in duplicate. Assay information for each pathogen target is available in supplementary table 3 [33]. Bacterial targets were *Aeromonas* spp.; *Escherichia coli* pathotypes including enteropathogenic (*EPEC* – *eae*, *bfpA*), enteroaggregative (*EAEC* – *aggR*, *aatA*, *aaiC*), enterohemorrhagic (*E. coli* O157), enterotoxigenic (*ETEC* – *LT*, *STp*, *STh*), Shiga toxin–producing (*STEC* – *stx1*, *stx2*), and enteroinvasive (*ipaH*); *Vibrio cholerae* (*hylA*); *Salmonella enterica* (*ttr*); *Campylobacter jejuni* and *coli*; *Helicobacter pylori*; *Clostridioides difficile*; *Mycobacterium tuberculosis*; and *Leptospira* spp. Viral targets included adenovirus 40/41, rotavirus, adenovirus, astrovirus, norovirus (GI and GII), SARS-CoV-2 (*N1*), Zika virus, and Human Immunodeficiency Virus (HIV). Protozoan targets included *Cryptosporidium* spp., *Giardia* spp., and *Entamoeba histolytica*. Helminth targets were *Ascaris lumbricoides*, *Trichuris trichiura*, *Ancylostoma duodenale*, and *Necator americanus.* Antimicrobial resistance was tracked via Class 1 integron (*intl1*). Fecal source tracking markers included mitochondrial DNA (human, poultry, canine), *Toxocara* spp., and avian 16S rRNA.

All cycling conditions for the RT-PCR were as follows: Initial hold step at 45°C for 20 minutes, followed by hold step at 95C for 10 minutes. In the PCR Stage, there were 50 cycles cycling between 95C for 15 seconds and 60C for 60 seconds. All steps also included a 1C/s heat increase/decrease. Two plates were prepared at a time and run on two separate QuantStudio 7 Flex RT-PCR machines. One included a PCR negative control, and the other contained a PCR positive control. Cq values (cycle threshold) were determined on each plate using manual thresholding.

### Mapping and Statistical Analysis Methods

Summary statistical analysis was completed using R-studio (R version 4.4.1) for absence/presence of all enteric pathogens using the manual thresholding and stratified by soil conditions, shade, and rainfall (CITE). A Cq cutoff of 35 cycles was used. A Poisson regression model with robust standard errors was fit to assess the association between predictor variables (sanitation infrastructure and land usage, wet soil, sun exposure, season) and our outcome variable (number of unique pathogens detected on TAC). The Poisson regression model was then analyzed for adjusted risk ratios (aRR), bounded by a 95% confidence interval [34, 35].

Mapping with ArcGIS-Pro was then used to visualize the number of unique pathogen detections across the sampling area of Maputo, Mozambique and compare against the field-team defined boundaries that separate non-sewered and Sewered System neighborhoods [28, 36]. Spatial clusters of unique pathogen detections were then visualized within the 4 different sampling locations (farm basin soils, sewered system areas, storm drainage areas, and the 150-meter buffer area).

## Analysis and Results

### Fecal Indicator Bacteria

*E. coli* concentrations were lower in farm basin soils (mean = 8.7×10^1^ per gram of soil, p < .001) than the non-sewered area (mean = 3.14×10^5^ per gram of soil), sewered area (mean = 1.23×10^3^ per gram of soil), and 150-meter buffer area (mean = 4.85×10^4^ per gram of soil). This was logarithmically distributed (figure 1).

**Figure 1:**
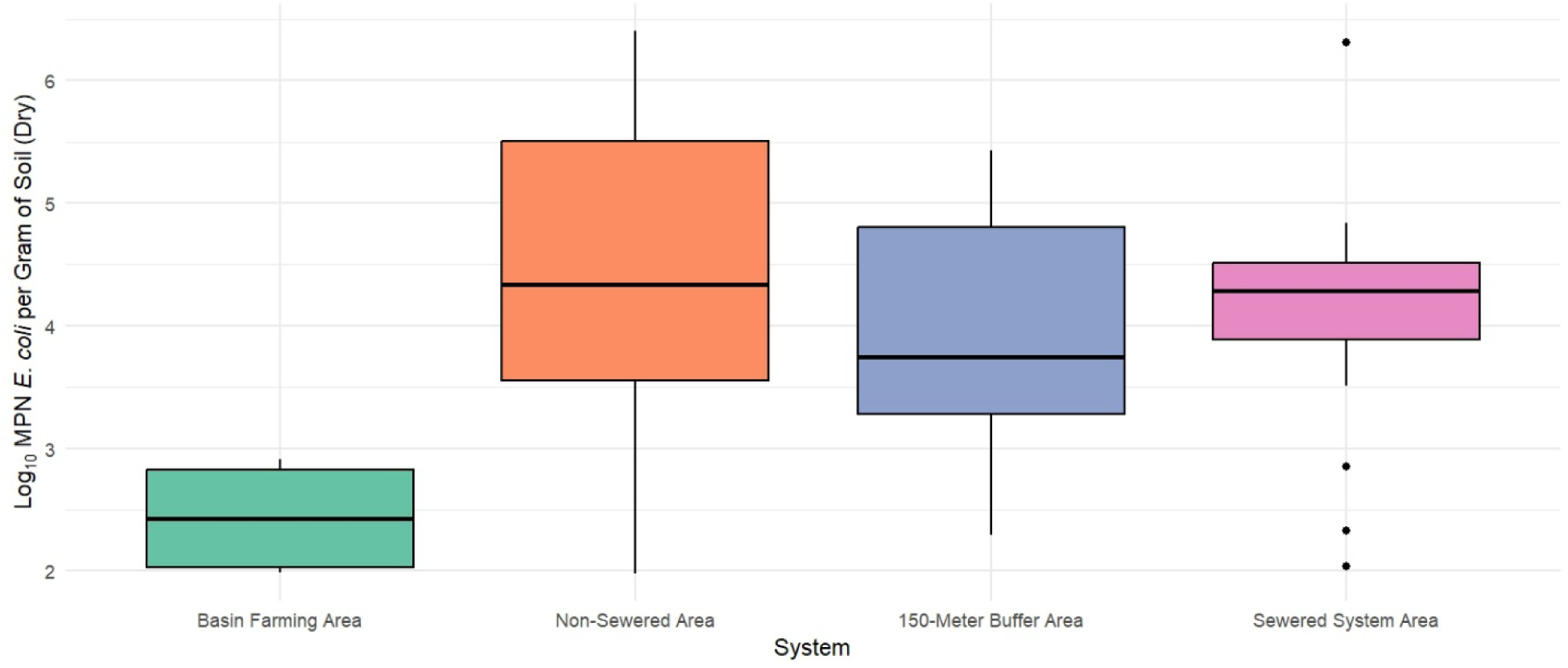
Box-And-Whisker plot of Log_10_ Most Probable Number (MPN) E. *coli* per gram of soil (dry) by sampling location. Basin farming area (n=19), non-sewered area (n=32), 150-meter buffer area (n=10), and sewered system area (n=20). The non-sewered area (mean = 3.14×10^5^ per gram), sewered area (mean = 1.23×10^3^ per gram), and 150-meter buffer area (mean = 4.85×10^4^ per gram) all have a significantly higher amount of *E. Coli* per gram of sample when compared to the basin farming area (mean = 8.7×10^1^ per gram) (p<.001).

### Unique Enteric Pathogen Detection Results

The three most common pathogens detected across all samples were *Aeromonas* (mean = 63.0%, CI95% = 52.3-72.7%), *Vibrio* hylA (mean = 51.1%, CI95% = 40.5-61.6%), and *Cryptosporidium* spp. (mean = 53.3%, CI95% = 42.6-63.6%) (Table 1). EAEC was found to be more common in non-sewered area (53.0%) and 150-meter buffer area (35.7%) when compared to the sewered area (26.7%). The other *E. coli* groups (EIEC, EPEC, ETEC, STEC) were found to have similar relationships across the public waste bin location groups as well. This trend was also seen within protozoan species, where *Cryptosporidium* had a higher prevalence in non-sewered area (66.7%) and the 150-meter buffer area (71.4%) when compared to the sewered area (20.0%) as well. The same was observed in *Giardia*, with higher prevalence in non-sewered area (25.6%) and the 150-meter buffer area (28.6%) when compared to the sewered area (15.0%). There was inconsistent detection of viruses across the viral pathogen group, but for adenovirus 40/41 and rotavirus there was a higher prevalence in non-sewered area (15.4%) and the 150-meter buffer area (14.3%) when compared to the sewered area (2.5%) as well. This trend was not present in some helminth species, where *Ascaris* had similar prevalence in the sewered (30%) and non-sewered areas (28.2%), but a higher prevalence in the 150-meter buffer area (42.9%).

**Table 1:** Average Percent Detection of Unique Enteric Pathogen, Fecal Source Trackers, and AMR by Location. Average Percent Detection of Unique Enteric Pathogens and Fecal Source Trackers by Location. This table shows average percent positive detection for bacteria, virus, protozoa, and helminth species within the 81 soil samples (10% in duplicate) in Maputo, Mozambique. Abbreviations used in the table are as follows: EAEC (Enteroaggregative *Escherichia coli)*, EIEC (Enteroinvasive *Escherichia coli),* EPEC (Enteropathogenic *Escherichia coli),* ETEC (Enterotoxigenic *Escherichia coli*), STEC (Shiga toxin–producing *Escherichia coli)*, HIV (Human immunodeficiency virus).

| Pathogens | Basin Farming Area (%) | 150-Meter Buffer Area (%) | Sewered System Area (%) | Non-sewered Area (%) |
| --- | --- | --- | --- | --- |
| **Bacteria** |  |  |  |  |
| *Aeromonas* | 36.8 | 85.7 | 60.0 | 69.2 |
| *C. difficile* | 5.3 | 14.3 | 5.0 | 2.6 |
| *Campylobacter jejuni/coli* | 10.5 | 21.4 | 15.0 | 12.8 |
| *Vibrio* | 10.5 | 64.3 | 45.0 | 69.2 |
| *E. Coli* | 10.5 | 35.7 | 20.0 | 51.3 |
| EAEC | 17.6 | 35.7 | 26.7 | 53.0 |
| EIEC | 15.8 | 35.7 | 20.0 | 25.6 |
| EPEC | 15.8 | 46.4 | 32.5 | 53.9 |
| ETEC | 12.3 | 31.0 | 8.3 | 35.1 |
| *Helicobacter pylori* | 15.8 | 7.1 | 5.0 | 2.6 |
| *Leptospira* | 10.5 | 0.0 | 5.0 | 2.6 |
| *Mycoplasma tuberculosis* | 21.1 | 28.6 | 10.0 | 2.6 |
| *Salmonella* | 5.3 | 21.4 | 20.0 | 23.1 |
| STEC | 5.3 | 21.4 | 10.0 | 14.1 |
| **Helminth** |  |  |  |  |
| *Ancylostoma duodenale* | 0.0 | 14.3 | 10.0 | 0.0 |
| *Ascaris lumbricoides* | 5.3 | 42.9 | 30.0 | 28.2 |
| *Necator americanus* | 10.5 | 7.1 | 5.0 | 0.0 |
| *Toxocara canis/cati* | 10.5 | 0.0 | 0.0 | 2.6 |
| *Trichuris trichiura* | 0.0 | 28.6 | 10.0 | 17.9 |
| **Protozoa** |  |  |  |  |
| *Cryptosporidium* spp. | 47.4 | 71.4 | 20.0 | 66.7 |
| *Entamoeba histolytica* | 15.8 | 7.1 | 5.0 | 2.6 |
| *Giardia* spp. | 5.3 | 28.6 | 15.0 | 25.6 |
| **Virus** |  |  |  |  |
| adenovirus 40/41 | 15.8 | 14.3 | 5.0 | 15.4 |
| astrovirus | 5.3 | 14.3 | 0.0 | 10.3 |
| HIV | 5.3 | 0.0 | 0.0 | 0.0 |
| norovirus | 0.0 | 14.3 | 0.0 | 0.0 |
| rotavirus | 5.3 | 14.3 | 0.0 | 15.4 |
| sapovirus | 0.0 | 28.6 | 5.0 | 10.3 |
| SARS-COV-2 | 15.8 | 7.1 | 5.0 | 2.6 |
| zika virus | 0.0 | 7.1 | 0.0 | 2.6 |
| Human mtDNA | 63.2 | 92.9 | 90.0 | 84.6 |
| Avian Bacteroides | 15.8 | 42.9 | 10.0 | 46.2 |
| Chicken mtDNA | 15.8 | 64.3 | 90.0 | 71.8 |
| Canine mtDNA | 5.3 | 57.1 | 70.0 | 66.7 |
| *Class 1 Integron-Integrase Gene* | 73.7 | 92.9 | 80.0 | 79.5 |

Cumulatively, viral pathogens were detected an average 5.7% of the time across all location groups, which was lower than unique bacterial pathogens (27.8%), protozoan pathogens (25.5%), and helminths (10.1%). The AMR gene target, Intl1, was detected within 80.4% of samples (CI95% = 70.6-87.8%) samples. Fecal-source tracking (FST) markers were positive for humans, canines, and poultry. FST markers for humans (87.6%), canines (66.2%), and poultry (76.5%) were detected in most public waste bin soil samples.

Shown in the box plot below in figure 2, the 150-Meter buffer sone area (mean = 10.5, median = 11.0, standard deviation = 3.3) and non-sewered areas (mean = 7.9, median = 8.5, s.d. = 3.0) had higher mean numbers of unique pathogens detected compared to both the basin farm soils (mean = 3.8, median = 2.0, s.d. = 4.8) and the sewered system areas (mean = 4.6, median = 4.5, s.d. = 2.5).

**Figure 2:**
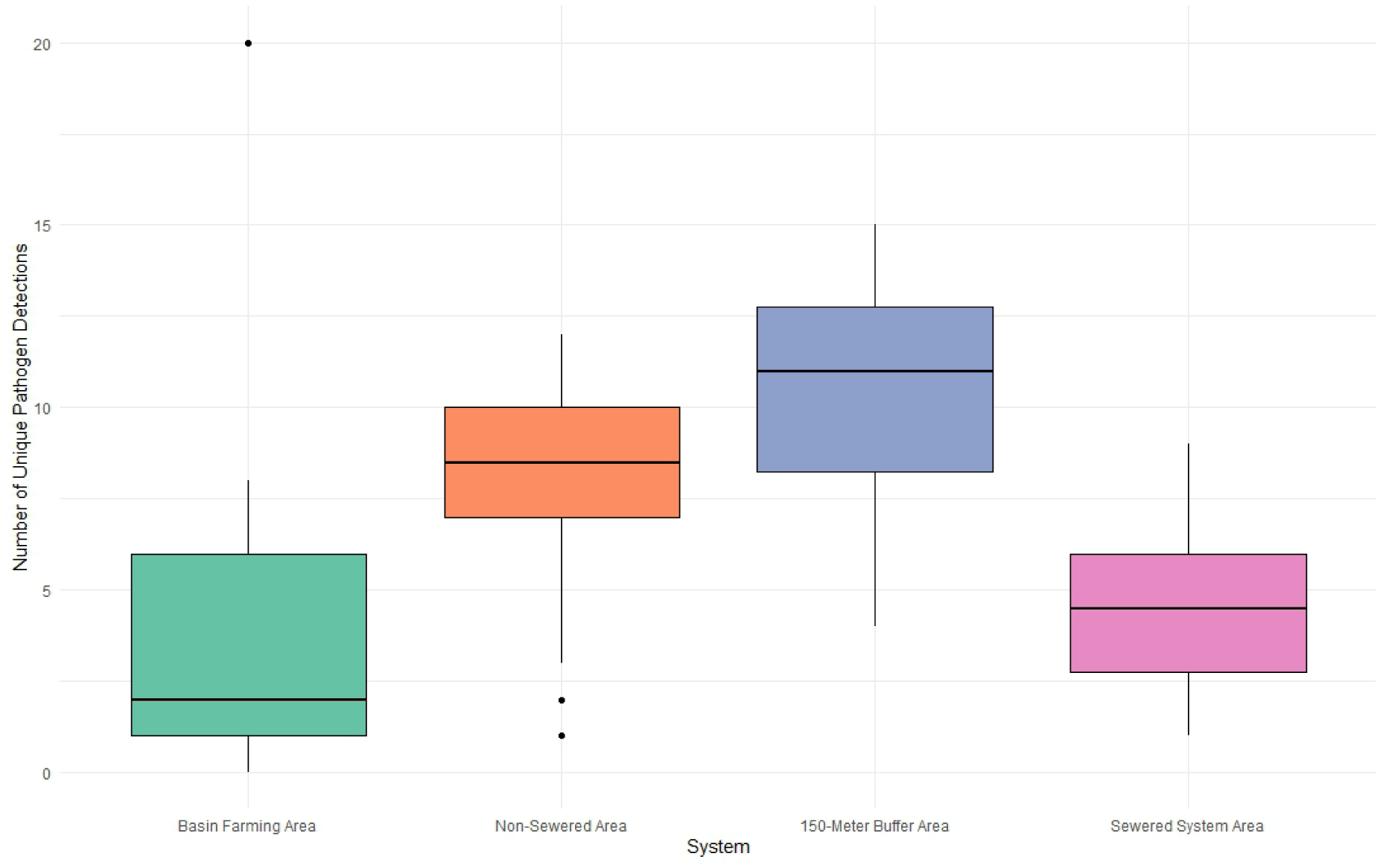
Box Plot of Unique Enteric Pathogen Detections by Public Waste Bin Location

Public waste bin soils from the non-sewered areas (aRR = 1.71 times greater, CI95% = 1.35-2.18, p<.001) and the 150-meter buffer area (aRR = 2.28 times greater, CI95%= 1.73-3.02, p<.001) were associated with an increased number of unique enteric pathogens detected compared to soil from the sewered system area (Table 2). There was no significant difference in risk for unique enteric pathogen detection between basin farm plot samples and sewered-area samples (p = .16). There was a not a statistically significant greater mean number of unique enteric pathogen detections in soil samples taken at public waste bins that were in complete shade when compared to those taken in complete sunlight (aRR = 1.18, CI95% = 0.96-1.46, p = .12). No statistically significant differences were found for the soil moisture or for the sampling period that the sample was collected (aRR = 1.10 times greater, CI95% = 0.92-1.30, p = 0.30).

**Table 2:** Poisson Regression Model Adjusted Risk Ratio (aRR) with 95% Confidence Interval.

| Predictor | Reference | aRR (95% CI) | p-value |
| --- | --- | --- | --- |
| Basin farm area | Sewered system area | 0.82 (0.60, 1.12) | 0.16 |
| Non-sewered area |  | 1.71 (1.35, 2.18) | <.001 |
| 150-Meter Buffer area |  | 2.28 (1.73, 3.02) | <.001 |
| Partial Sun | Complete Sun | 1.08 (0.88, 1.33) | 0.45 |
| Complete Shade |  | 1.18 (0.96, 1.46) | 0.12 |
| Dry | Wet | 1.10 (0.92, 1.30) | 0.30 |

### Mapping Analysis

The 150-meter buffer area around the boundary between the non-sewered and the sewered areas had a greater density of high unique enteric pathogen detection sites when compared to the sites in the non-sewered, sewered, and basin farming areas. Through ArcGIS Pro, this boundary also showed to be a transitionary zone between grid style neighborhoods of the Southeast part of Maputo where there are sewer systems and the North and Northeast parts that are non-sewered areas with storm drainage systems.

Mapping the site locations also indicated that sewered system public bin soil samples in the Southeast part of the city had the lowest number of unique enteric pathogen detections when compared to the urban public waste bin soil samples. These sewered area public bin soils had a reduced number of unique enteric pathogen detections the further the distance they were from the 150-meter buffer area.

Overall, mapping the public waste bin soils indicated that non-sewered areas had the highest number of unique enteric pathogen detections in public waste bins in Maputo and the Southeast part of Maputo had a lower number of unique enteric pathogen detections.

**Figure 3:**
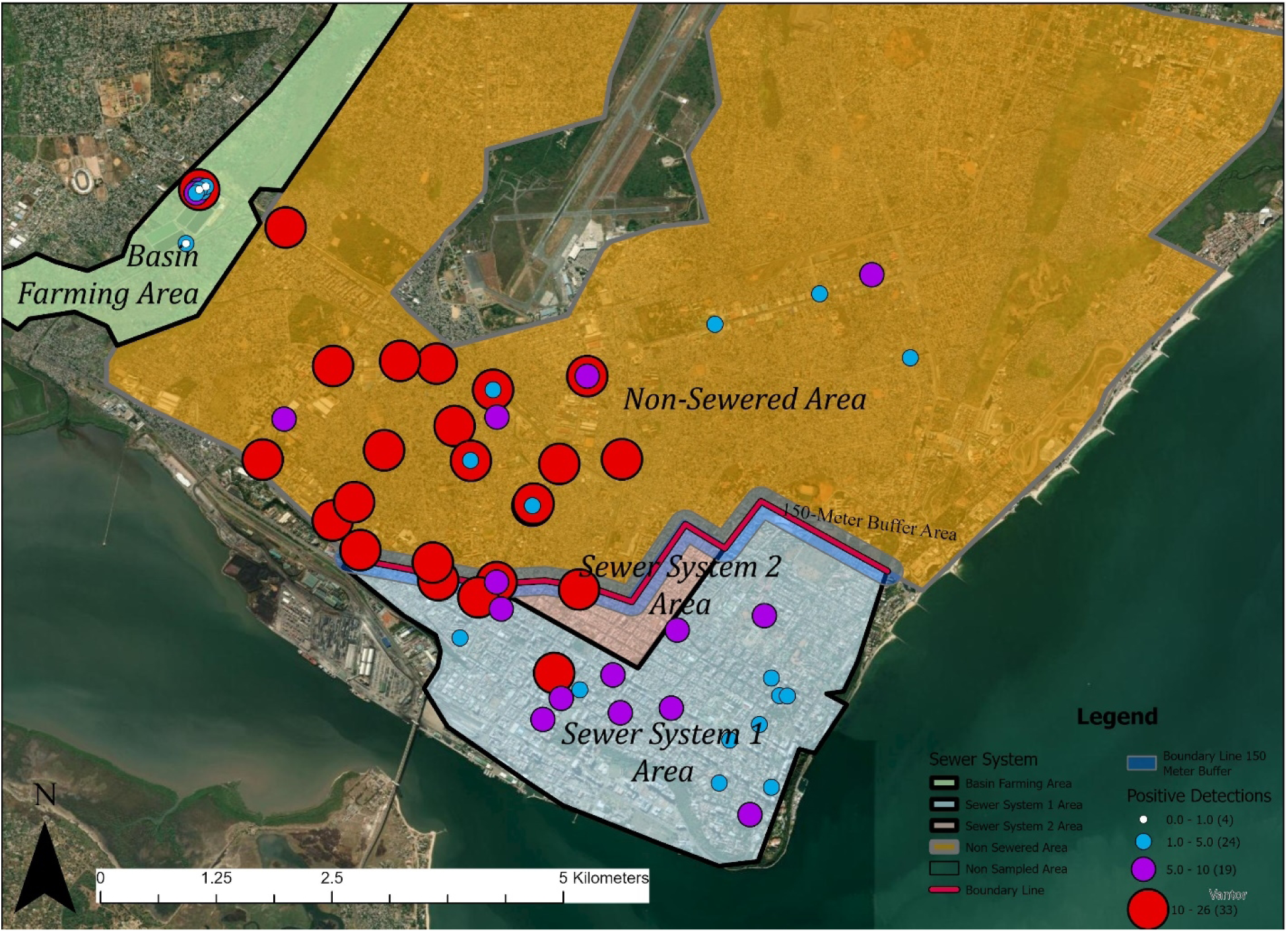
Unique Pathogen Detection by Soil Location in Maputo, Mozambique. The map shows the average number of unique enteric pathogen detection across the sampling area in Maputo, Mozambique. Positive detections are shown in circles, sized and colored by the average number of unique pathogen detections. Red circles indicate sample sites that had 10 or more unique enteric pathogen detections. Purple circles indicate between 5-10 detections, blue circles indicate 1-5 detections, and white circles indicate 0 detections. The basin farm sampling locations are shown in the green Northwest quadrant of the map outside of the urban area. The non-sewered area areas are defined by the translucent orange polygon. The sewered system areas are shown in translucent light-blue and red polygons in the Southeast part of the map. The 150-meter buffer area is shown by the dark-blue translucent polygon with the red line centering it.

## Discussion

We observed widespread fecal contamination in soil adjacent to public waste bins in Maputo, Mozambique. Local sanitation infrastructure was a predictor of the number of unique enteric pathogens in these soils. While we did not compare our results against the local enteric infection burden, our results indicate the soils adjacent to public waste bins are sensitive to variations in sanitation infrastructure. Unlike wastewater, which is only present in areas covered by piped sewerage systems, fecally contaminated soils are often present in sewered and unsewered areas and represent an alternative matrix for WBE in low resource settings.

The design of sanitation infrastructure may explain why we observed fewer enteric pathogens in soil adjacent to public waste bins in the sewered area compared to the non-sewered area. Piped sewage systems convey fecal waste away for offsite treatment or discharge, while onsite systems store fecal wastes in a pit or tank until the waste is emptied and either buried onsite or transported offsite for disposal [30]. Offsite conveyance by a sewer system reduces the amount of fecal waste present in an area that could be disposed of as solid waste, and may be limited to bagged animal feces, diapers and other materials soiled with feces including anal cleansing materials. However, in non-sewered areas there is a greater volume of fecal waste present that could be plausibly disposed of via solid waste, including fecal sludge, diapers, bagged human or animal fecal waste, open defecation, and other materials soiled with feces.

The sewered area, located in KaMpfumo district, was extensively developed during the colonial period and followed conventional European urban planning principles intended to serve the Portuguese colonial rulers. In contrast, the non-sewered areas, Nhlamankulu and KaMaxakeni districts, evolved more informally and historically received significantly less investment compared to KaMpfumo [37, 38]. In the post-colonial period, the sewered area within Maputo remains the central business district as well as containing most of the higher income housing in the city. The non-sewered areas remain as the poorer, more populated districts of Maputo [38]. This relationship between the two is evidenced by the historically higher counts of poverty in the non-sewered areas when compared to the sewered areas by the World Bank and Mozambican Censuses between 1997-2017 [31, 36]. In visits to Maputo in 2022 for sampling, evidence of this relationship to remain in place and may be a driver for pathogen exposure in both the domestic and public domains.

The 150-meter buffer area had the highest average of unique enteric pathogen detections compared with other sample locations. This area blends civil infrastructure of both the sewered and non-sewered areas as described above. We posit that the higher detections within this area are likely due to lack of sewerage coverage and higher pedestrian traffic observed. Although correlations between pedestrian traffic and solid waste or open defecation were not examined as part of this study, the central location of the buffer area would agree with our hypothesis that these areas received larger amounts of solid, human and non-human waste.

Previous research on solid waste exposure has indicated that access to both municipal waste collection and basic sanitation are needed to prevent solid waste sites such as public waste bins from becoming reservoirs for pathogens [39]. Within Maputo, lack of emptying and responsibility over emptying has also contributed, meaning that waste can remain within poorer areas for longer [38, 40]. Two empirical studies have found a positive correlation with distance and size of solid waste at public waste bins with increasing the risk of pathogen prevalence [41, 42]. In a city like Maputo, where access to bins and public toilets is limited, this can mean that public waste bins and the soils around them can act as reservoirs for pathogens.

In fact, Mittal et al. found that walking was the preferred or only available method of transportation for many people within Maputo and that people that lived within poorer areas were more likely to travel further distances [43]. This supports the heavier foot traffic observed within the 150-meter buffer zone, and additional possible driver for the increased unique pathogen detections found there. Although our study did not assess proximity or emptying frequency of solid waste as exposure risks, the increased number of unique enteric detections in highly trafficked areas concurs with the conclusions drawn within these prior studies that lack of access to basic sanitation and the undercapacity of the municipal solid waste department has potentially resulted in increases of human waste around public waste bins that represent an environmental hazard to pedestrians and sanitation workers.

Recent research has also underscored the public health risks associated with mixing of unmanaged solid and human waste in the global south [44, 45]. Open and improper waste dumping contaminates soil and water with enteric pathogens, which can result in chronic and acute health issues such as respiratory diseases, gastrointestinal infections, and adverse neonatal outcomes, although this remains unstudied within this context [26, 29, 46, 47]. This prior research underpins the risks apparent within the public domain in Maputo, where the findings of this study indicate this is happening within both sewered system and non-sewered areas. As pathogen detections averaged between 4.6 and 10.5 unique pathogens per soil sample at public waste bins, there is an apparent need to assess the public health risk associated with public waste bin usage, especially within heavily trafficked areas.

As these public waste bin locations are at street level, they also pose a risk of exposing neighborhoods further during heavy rainfall and flooding events or reversely at risk of being exposed during these failure events to non-sewered waste from the domestic domain [48-50]. Capone et al. (2025) found a minor increase in pooled pathogen concentration across environmental sampling matrices during heavier rain fall periods [28], further evidencing this potential risk. The relationship between the domestic and public domain in this way may be circular; a positive feedback loop can occur where pathogens exist in both domains and can travel between each domain through foot-traffic, improper management of waste, or flood events.

The combined use of public waste bins soils as a matrix of study and qPCR as a tool of analysis for unique pathogen detections could be beneficial for monitoring pathogen flow within complex urban systems such as Maputo [51, 52]. Similar unique pathogens were detected within the domestic and public domain in earlier portions of the MapSan trials and environmental pathogen monitoring studies in Maputo [27, 28] as well, where soils samples can be taken not only in both domains, but also within both sewered and non-sewered areas. Thus, this matrix allows for direct comparison, whereas other matrices such as fecal sludge and wastewater are not directly comparable.

This study was completed using convenience sampling and may not be representative of all public waste bins across the sewered and non-sewered areas within Maputo. Although seasonality was initially assessed through two sampling periods, insignificant difference between periods and between samples taken on rainy or dry days was found. A limitation of a qPCR-based approach is that it diminishes understanding of pathogen viability within the soil. Additional study would be needed to prove a connection between seasonality or rainfall events and unique pathogen detection and better observational data would be needed to assess how frequently these sample locations are inundated with water during rainfall events. Additionally, this study did not count the number of instances that fecal matter was observed at site, but we observed their presence at the sites during sampling. This included diapers, human waste, animal waste, as all are possible sources for the spread of enteric pathogens within the urban environment. Fecal source tracking markers verified these visual observations where fecal-source trackers for humans, chickens, and dogs were detected within the majority of soils adjacent to public waste bins but infrequently from soils at subsistence farms (S3).

The results of this study indicated that there was a risk of increased number of unique pathogens in non-sewered areas, as well as sewered areas within a 150-meter distance of these areas when compared to sewered areas in the Southeast part of Maputo. These areas pose a risk of continuing the spread of enteric pathogens between domestic and public domains. The findings also indicate the potential health risks for public bin users and sanitation workers responsible for removal of solid waste, as well as the potential role of those who traffic these areas as spreaders of enteric pathogens from public to domestic domain and vice versa.

Soil samples appear to be sensitive to the variation in local sanitation infrastructure and represent an alternative matrix that could potentially be used for environmental surveillance as a component of WBE programs. Thus, the findings of this study emphasize the importance of diversifying matrices of environmental surveillance within urban environments and developing unique sampling strategies as the context requires.

## Data Availability

Protocols and data used in this paper are available at a dedicated data repository at Open Science Framework (OSF.io). Permanent link: https://osf.io/h5epg/.

## Appendix A: Supplemental Data Tables

**Table S1:** Summary Statistics for Unique Enteric Pathogen Detections by Site Location.

| *Site Location* | *Mean* | *Median* | *Std Dev* | *Min* | *Max* |
| --- | --- | --- | --- | --- | --- |
| Basin farm area (n=19) | 3.80 | 2.00 | 4.77 | 0 | 20 |
| Non-sewered area (n=32) | 7.90 | 8.50 | 3.04 | 1 | 12 |
| Sewered system area (n=20) | 4.60 | 4.50 | 2.48 | 1 | 9 |
| 150-Meter Buffer Area (n=10) | 10.5 | 11.00 | 3.31 | 4 | 15 |

**Table S2:** Poisson Regression Model Adjusted Risk Ratio (aRR) with 95% Confidence Interval.

| *Predictor* | *Reference* | *aRR (95% CI)* | *p-value* |
| --- | --- | --- | --- |
| Basin farm area | Sewered system area | 0.82 (0.60, 1.12) | 0.16 |
| Non-sewered area |  | 1.71 (1.35, 2.18) | <.001 |
| 150-Meter Buffer area |  | 2.28 (1.73, 3.02) | <.001 |
| Partial Sun | Complete Sun | 1.08 (0.88, 1.33) | 0.45 |
| Complete Shade |  | 1.18 (0.96, 1.46) | 0.12 |
| Dry | Wet | 1.10 (0.92, 1.30) | 0.30 |

**Table S3:**
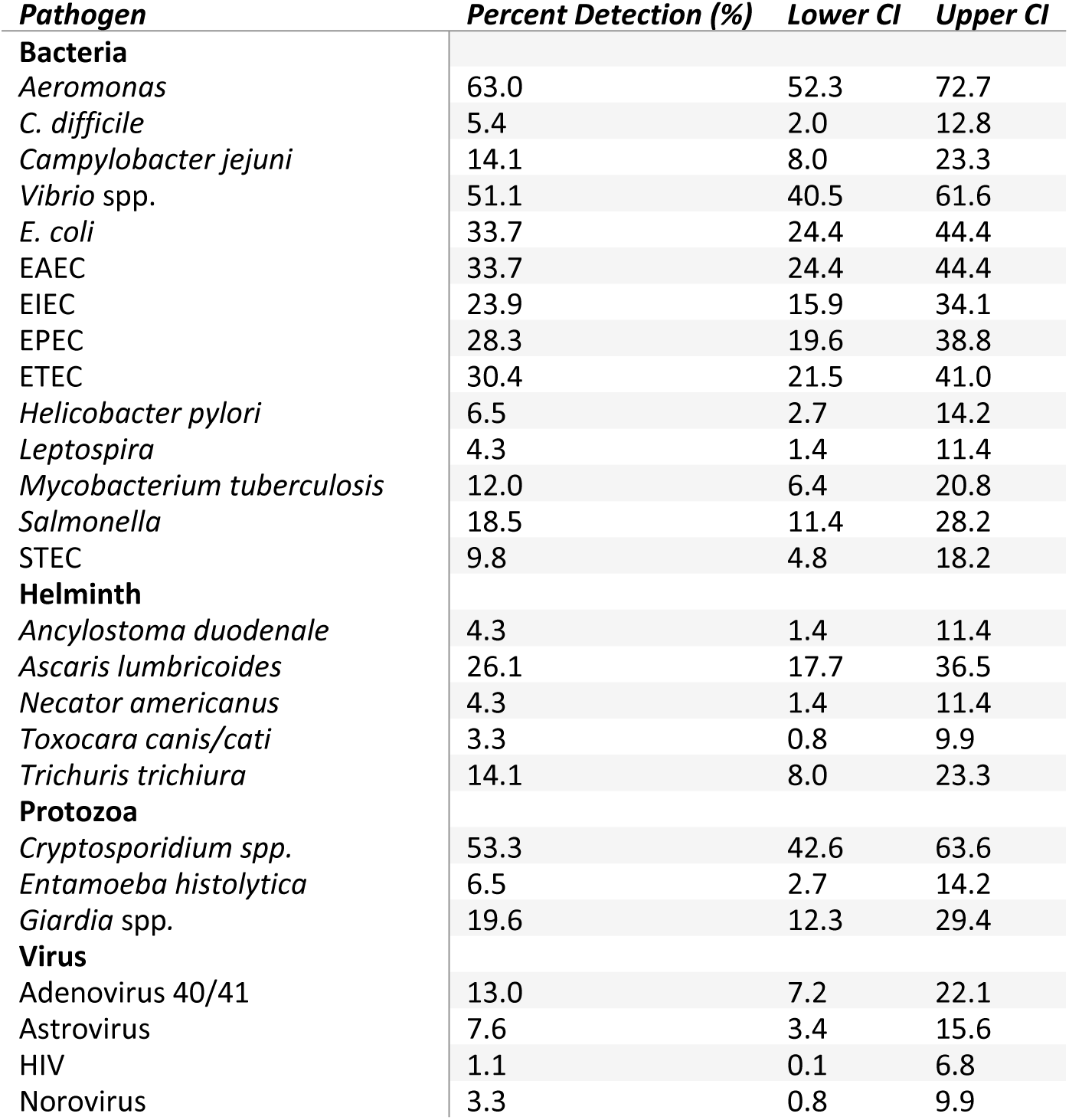
Target Percent Detection with 95% Confidence Interval.

**Table S4:**
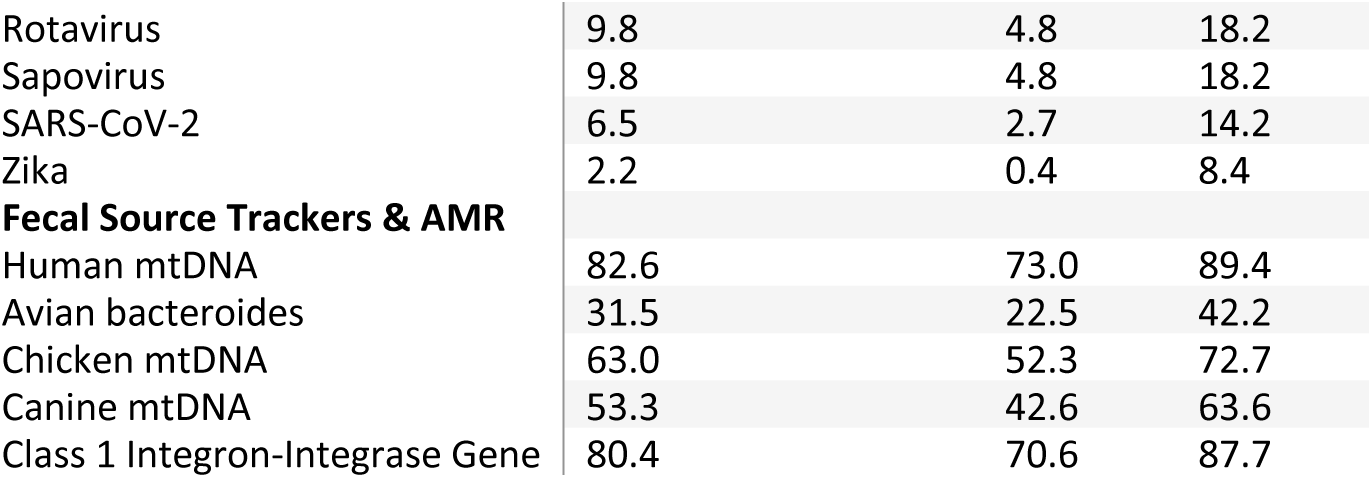
TAC Assay Information (See Supplemental File)

## Notes

### Competing Interest Statement

The authors have declared no competing interest.

### Funding Statement

This study was funded by the Bill & Melinda Gates Foundation (www.gatesfoundation.org) Grant OPP1137224. The funders had no role in study design, data collection and analysis, decision to publish, or preparation of the manuscript. D.C. was supported in part by an NIH T32 Fellowship (5T32ES007018-44).

### Author Declarations

This work was excempt from IRB approval.

